# Postoperative morbidity after liver resection- A Systemic review, meta-analysis, and metaregression of factors affecting them

**DOI:** 10.1101/2021.04.06.21254984

**Authors:** Bhavin Vasavada, Hardik Patel

## Abstract

**Aim of the study:** This systemic review and meta-analysis aimed to analyze post-operative morbidity after liver resection, and also study various factors associated with mortality via metaregression analysis.

**Material and Methods:** PubMed, Cochrane Library, Embase, google scholar, web of science with keywords like ‘liver resection”; “mortality”;” hepatectomy”. Weighted percentage post-operative morbidities were analyzed. Meta-analysis and meta-regression were done by the DerSimonian-Liard random effect model. Heterogeneity was assessed using the Higgins I2 test. Publication bias was assessed using a funnel plot. Funnel plot asymmetry was evaluated by Egger’s test. Morbidity was defined as any postoperative morbidity mentioned.

**Results:** A total of 46 studies was included in the final analysis. Total 45771 patients underwent liver resections. 16111 patients experienced complications during the postoperative period. Weighted post-operative morbidity was 30.2% (95 % C.I. 24.8-35.7%). Heterogeneity was high with I^2^ 99.46% and p-value <0.01. On univariate analysis, major liver resections were significantly associated with heterogeneity. (p=0.024). However, residual heterogeneity was still high with I^2^ 98.62%, p<0.001. So, multifactor metaregression analysis major hepatectomy (p<0.001), Open hepatectomy (p=0.001), cirrhotic liver (p=0.002), age (p<0.001), blood loss (p<0.001), and colorectal metastasis (p<0.001) independently associated with postoperative morbidity. Residual heterogeneity was moderate I2= 39.9% and nonsignificant p=0.189.

**Conclusion:** Liver resection is associated with high postoperative morbidity and various factors like major hepatectomy, Open hepatectomy, cirrhotic liver, blood loss, and colorectal metastasis were associated with morbidity and responsible for heterogeneity across the studies.

## Background

Liver resection is now established curative treatment for various malignant and benign liver pathology. Currently, Hepatocellular carcinoma and colorectal liver metastasis are the most common indications for liver surgery as well as primary and secondary liver malignancies. [1,2].

Perioperative care and surgical techniques for liver resections have improved significantly in the past few years, resulting in improved perioperative outcomes. Despite improved outcomes liver resection is still associated with very high morbidity and mortality. Also, there is a lack of uniformity in reporting the outcomes across various centers. [3,4,5]. Indications also vary with Europe having a predominance of colorectal cancer metastasis as indications whereas in Asia hepatocellular carcinoma is the most common indication for liver resection. [6].

This systemic review and meta-analysis aimed to analyze any post-operative morbidity after liver resection, and study various factors associated with mortality via univariate and multivariate metaregression.

## Material and Methods

The study was conducted according to the Preferred Reporting Items for Systematic Reviews and Meta-Analyses (PRISMA) statement (2020) and MOOSE guidelines. [7,8]. We registered our protocol in the PROSPERO database [246263]. We conducted a literature search as described by Gossen et al. [9]. PubMed, Cochrane Library, Embase, google scholar, web of science with keywords like ‘liver resection”; “morbidity”;” hepatectomy”. Two independent authors extracted the data (B.V and H.P). We evaluated the last studies published in the last 5 years to see recent trends in 90 days mortality. In case of disagreements, decisions are reached on basis of discussions.

### Definitions

We defined morbidity as any postoperative complications according to Clavien-Dindo classifications grade 1-5. [10]

### Statistical Analysis

The meta-analysis was done using Review Manager 5.4 and the JASP Team (2020). JASP (Version 0.14.1)(University of Amsterdam). Weighted percentage morbidities with 95% confidence intervals were used. Univariate metaregression was done by DerSimonian-Laird methods. Major hepatectomy, Age, open surgery, cirrhotic livers, blood loss, hepatectomy for hepatocellular carcinoma, hepatectomy for colorectal liver metastasis were taken as covariates in metaregression analysis to study their association heterogeneity of the meta-\analysis. Factors with a p-value less than 0.05 were entered in the multivariate metaregression model and then we decided to check for residual heterogeneity, if residual heterogeneity is still significant, we decided to enter all co-variants in the multivariate model to look for mixed effects. Heterogeneity was assessed using the Higgins I^2^ test, with values of 25%, 50%, and 75% indicating low, moderate, and high degrees of heterogeneity, respectively, and assessed p-value for the significance of heterogeneity and tau2 and H2 value [11]. The random-effects model was used in meta-analysis. The random-effects model was used in meta-analysis.

### Assessment of Bias

Cohort studies were assessed for bias using the Newcastle-Ottawa Scale to assess for the risk of bias [12,13] Publication bias was assessed using a funnel plot. Funnel plot asymmetry was evaluated by Egger’s test.

### Inclusion and Exclusion criteria for studies

#### Inclusion criteria

- Studies with full texts
- Studies published in last 5 years.
- Studies mentioning post-operative morbidity rates.
- Studies which evaluated liver resections for different etiologies.
- English language studies.

#### Exclusion criteria

- Studies which were not fulfilling the above criteria.
- Duplicate studies.
- Studies which included analysis of liver resection for single etiology.

## Results

### Data extraction, study characteristics, and quality assessment

‘PUBMED’, ‘SCOPUS’, and ‘EMBASE’ databases were searched using keywords and the search strategy described above. Initially, 4256 studies published in the last 5 years were screened from the above search strategy, and 26 additional records were screened from references of the above studies. After removing duplicates 3348 studies screened again. 3265 studies excluded meeting the exclusion criteria. 83 full-text articles were evaluated. 37 full texts article removed as they did not mention post-operative morbidity rates.46 studies were included in the final qualitative and quantitative analysis. The risk of bias summary is mentioned in Figure 2. Study characteristics are mentioned in table 1. All except one study included are either retrospective studies or propensity score-matched analysis from retrospective data. In propensity score-matched analysis unmatched total data were analyzed for postoperative morbidity. Only one study was randomized control trial[51] in that also we analyzed total complications in both the group and total population. So, effectively all the data were retrospective.

**Table 1.** Study characteristics.

| STUDY | YE<br>R | total | MAJOR<br>HEPATECTOMY | ope<br>n | cirr<br>ho<br>sis | Age | blood<br>loss | hepatocellu<br>lar<br>carcinoma | colorectal<br>lm | total<br>morbidity |
| --- | --- | --- | --- | --- | --- | --- | --- | --- | --- | --- |
| al-saeedi | 2020 | 209 | 209 | 209 | 83 | 60 | 1500 | 16 | 59 | 63 |
| al-saif | 2019 | 111 | 48 | 89 |  | 52.6 |  | 14 | 1 | 40 |
| alam | 2016 | 77 | 45 | 77 |  | 49 |  | 11 | 35 | 14 |
| amico | 2016 | 84 | 38 | 84 | 6 | 54.0<br>1 |  | 9 | 30 | 35 |
| atili | 2019 | 10 | 10 | 8 | 0 | 60.3 |  | 3 | 0 | 11 |
| braunwart<br>h | 2019 | 458 | 242 | 430 | 26 | 61 | 400 | 49 | 149 | 17 |
| chin | 2020 | 472 | 56 | 472 | 168 | 62 | 682 | 309 | 163 | 21 |
| CHOPINE<br>T | 2018 | 75 | 75 | 75 | 75 | 60 | 209 | 75 | 0 | 538 |
| CIESLAK | 2016 | 163 | 163 | 163 | 103 | 63 | 211 | 12 | 0 | 43 |
| fujii | 2019 | 103 | 103 | 103 | 63 | 67 | 2500 |  |  | 237 |
| garnier | 2018 | 111 | 111 | 111 | 37 | 66 |  | 9 | 66 | 21 |
| gelli | 2018 | 1803 | 619 | 180<br>3 | 226 | 61 |  |  |  | 129 |
| Goh | 2017 | 195 | 12 | 0 | 50 | 60 | 200 | 102 | 46 | 73 |
| Grey | 2016 | 66 | 31 | 62 |  | 62 |  | 18 | 33 | 69 |
| gupta | 2018 | 26 | 12 | 26 |  | 82 | 400 | 4 | 20 | 65 |
| halls | 2017 | 2861 | 429 | 222 | 220 | 64 |  | 189 | 1630 | 10 |
| higashi | 2015 | 144 | 138 | 144 |  | 65.1 |  | 91 | 12 | 95 |
| hobeika | 2018 | 3150 | 80 | 0 | 774 | 62 | 350 | 1247 | 714 | 12 |
| lee | 2020 | 72 | 72 |  |  | 57 |  | 48 | 9 | 84 |
| lemke | 2017 | 749 | 516 | 681 | 25 | 64 |  | 56 | 489 | 71 |
| Levisandri | 2018 | 225 | 225 |  |  | 62.4 | 400 | 77 | 106 | 496 |
| Lin | 2019 | 281 |  | 232 |  | 55 | 300 |  |  | 30 |
| longbottho<br>m | 2019 | 283 | 283 | 283 | 124 | 62 |  | 36 | 237 | 27 |
| longbottho<br>m1 | 2019 | 49 | 49 | 49 | 20 | 78 |  |  |  | 453 |
| Matteo | 2017 | 340 | 48 | 340 |  | 65 | 300 | 82 | 196 | 353 |
| Ome | 2017 | 142 | 0 | 79 | 47 | 68 | 600 |  |  | 34 |
| Pietraz | 2018 | 195 | 47 | 0 |  | 68.2 |  | 22 | 130 | 78 |
| ratti | 2018 | 204 | 114 | 102 | 8 | 62 |  |  |  | 71 |
| ruzzenente | 2017 | 803 | 242 | 803 | 109 | 65.5<br>4 |  | 176 | 279 | 496 |
| saleema | 2017 | 75 | 30 | 75 | 30 | 52 | 665 | 37 | 10 | 30 |
| urbani | 2017 | 60 | 0 | 60 |  | 60 | 150 | 7 | 48 | 27 |
| vanmirelo | 2016 | 956 | 466 | 956 | 320 | 64 | 600 | 61 | 657 | 453 |
| vigano | 2020 | 2212 | 2212 | 2212 | 0 | 62 |  | 414 | 947 | 353 |
| worhunsy | 2016 | 167 | 30 | 0 | 48 | 58 | 100 | 56 | 63 | 34 |
| Yip | 2016 | 223 | 103 | 200 |  | 67 |  | 25 | 105 | 78 |
| Zadyfudiu<br>m | 2020 | 3530 | 1263 | 3530 | 668 | 60 |  |  |  | 503 |
| Xu | 2017 | 2008 |  |  |  |  |  |  |  | 292 |
| weinberg | 2019 | 58 | 58 | 58 |  | 66 |  |  |  | 27 |
| Uneo | 2017 | 90 | 58 | 90 |  | 70 | 360 | 57 |  | 19 |
| UCHIDA | 2017 | 95 |  | 95 | 53 | 70 | 132 | 71 | 12 | 17 |
| tran | 2016 | 5227 | 5227 | 5227 |  | 60 |  |  |  | 3131 |
| synder | 2019 | 3912 | 3912 | 3912 |  | 58 |  | 1332 | 1795 | 1378 |
| shwaartz | 2016 | 1005 | 1005 | 896 | 83 | 57 | 514 | 271 |  | 452 |
| ross | 2016 | 11934 | 4399 |  |  | 59 |  | 1837 | 4618 | 5364 |
| leroux | 2018 | 155 | 26 | 95 | 62 | 65 |  | 62 | 93 | 60 |
| hughes | 2016 | 603 | 336 | 570 |  | 60.5 |  |  |  | 207 |

**Figure 1.**
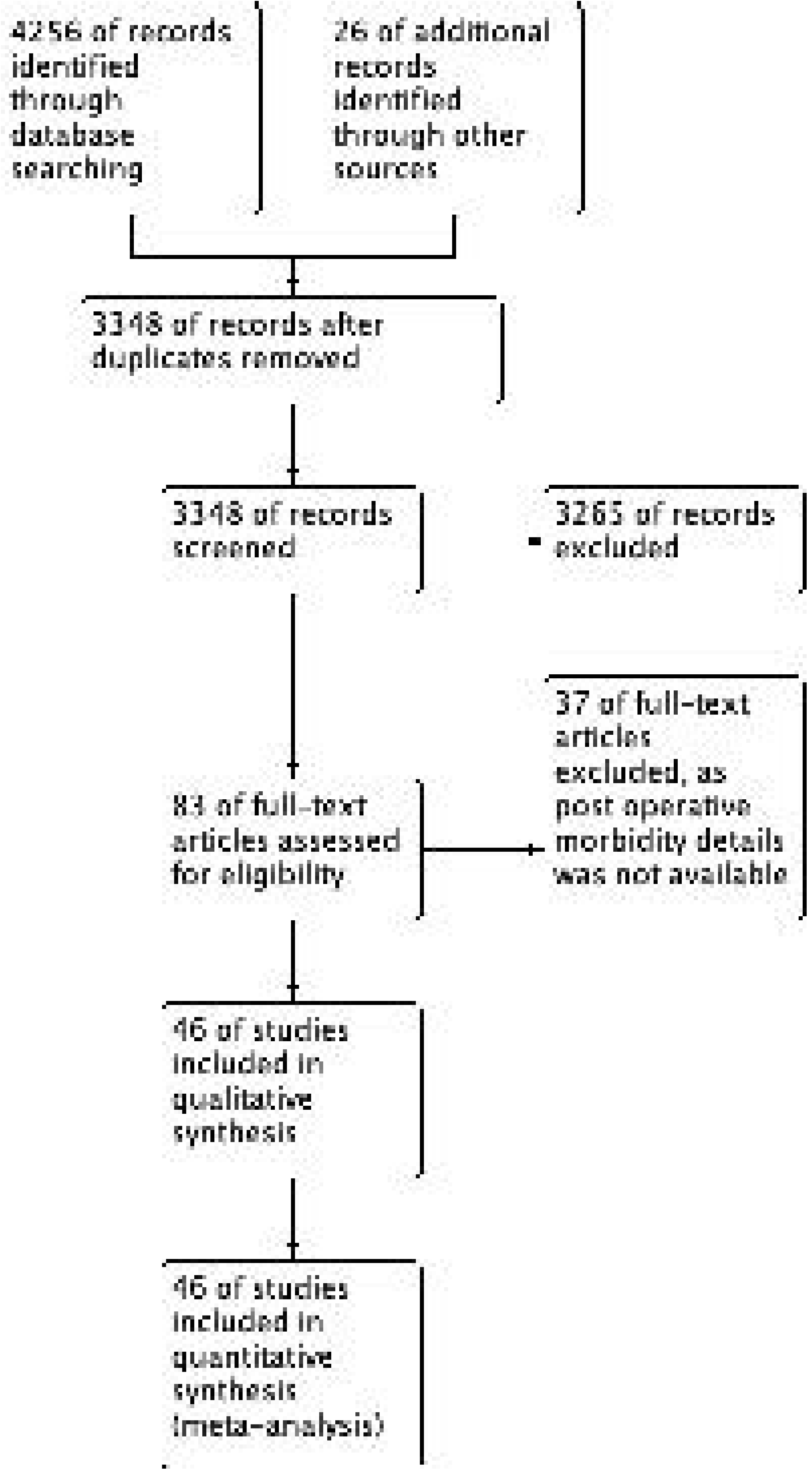
PRISMA flow diagram.

**Figure 2.**
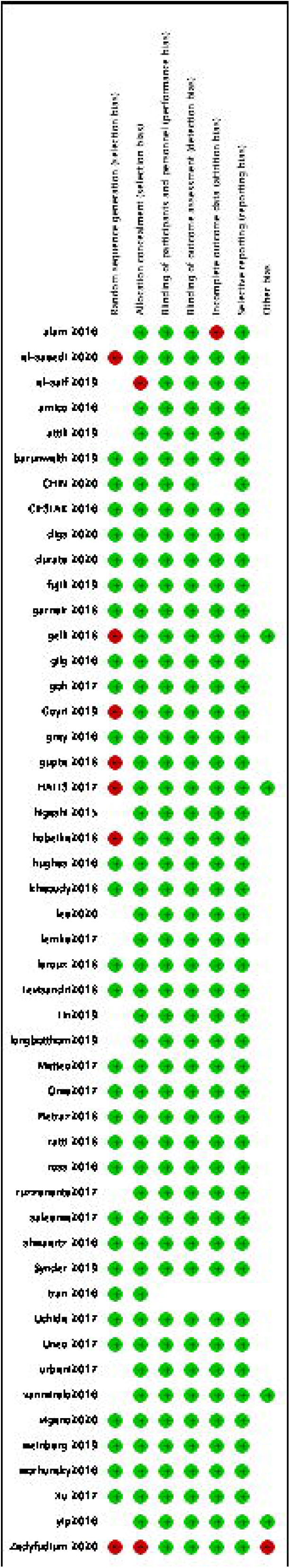
Risk of Bias Summary.

### Weighted post-operative morbidity

A total of 46 studies was included in the final analysis. [14-58]. Total 45771 patients underwent liver resections. 16111 patients experienced complications during the postoperative period. Weighted post-operative morbidity was 30.2% (95 % C.I. 24.8-35.7%). [Figure 3]. However, the heterogeneity of the analysis was high with I2 99.463%, tau2 0.034, and Q value of 8379.71, df 45. A p-value of heterogeneity was highly significant (p<0.001).

**Figure 3.**
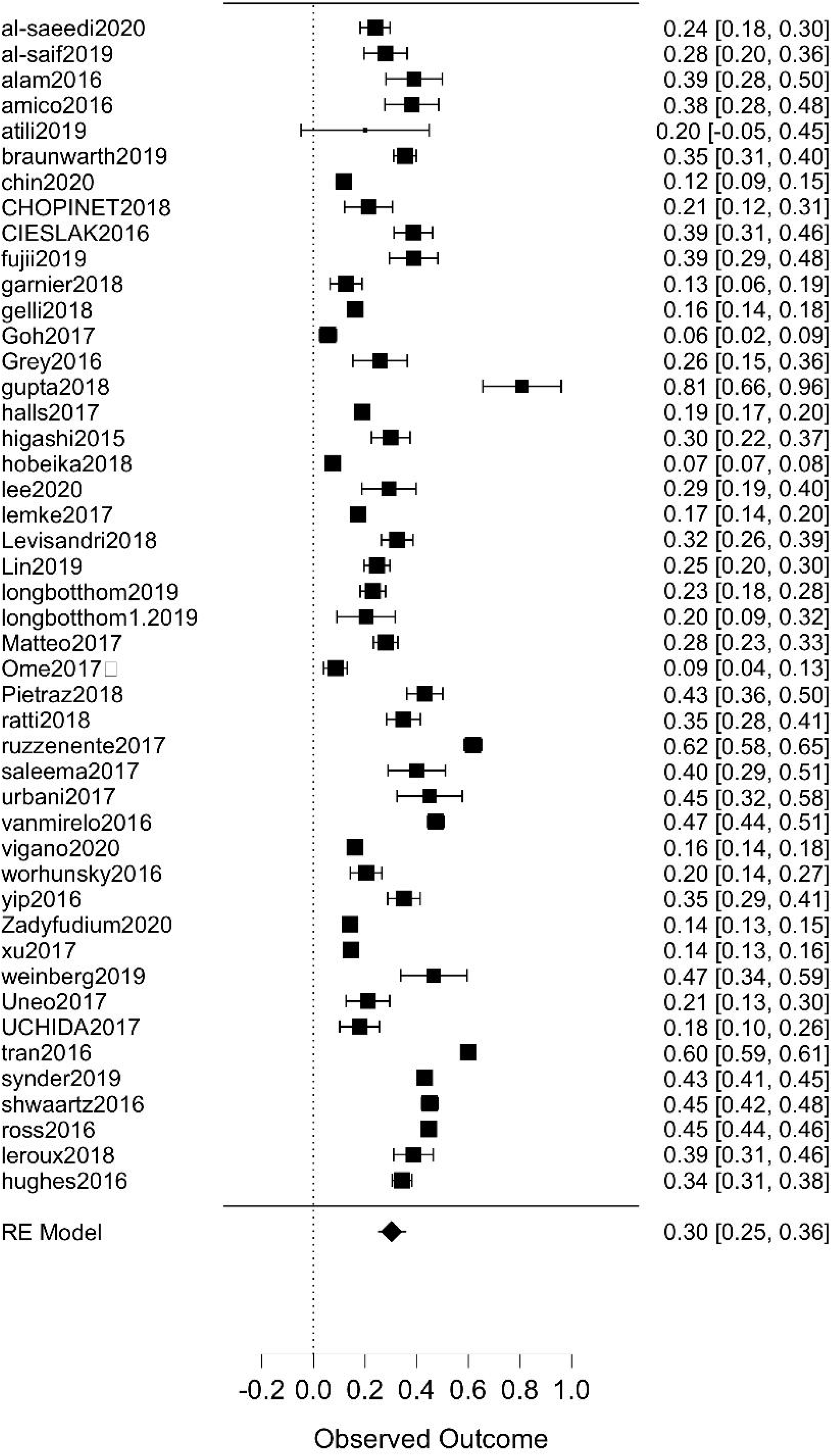
Forest plot of post-operative morbidity.

### Publication Bias

Figure 4 mentions the funnel plot for publication bias. Egger’s test showed nonsignificant publication bias with a p-value of 0.202.

**Figure 4.**
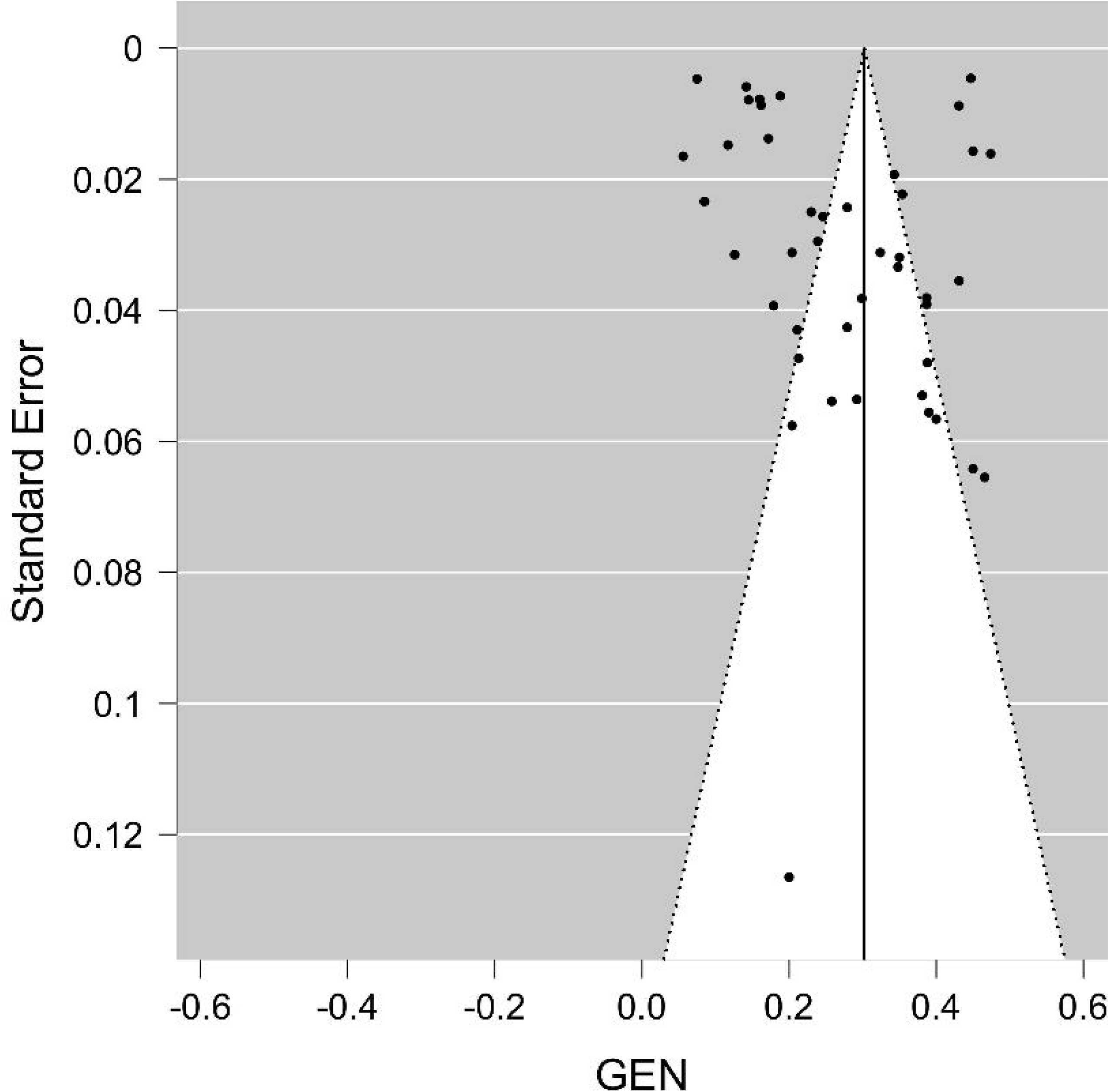
Publication bias for post-operative morbidity.

### Metaregression analysis

We analyzed various covariates like major hepatectomy, Age of the patient, blood loss, open surgery, liver resections were done for hepatocellular carcinoma or colorectal liver metastasis and cirrhotic liver to check for their association with heterogeneity in the analysis and hence postoperative morbidity. On univariate analysis, only major hepatectomy was significantly associated with heterogeneity with a p-value of 0.024. However, residual heterogeneity was still highly significant with I2 is 98.5%, Q value 2988.013, df 41, Tau2 value of 0.015. So, we entered all the factors in the multiple meta-regression model.

### Multiple metaregression model

On multiple metaregression model major hepatectomy (p<0.001), open surgery(p=0.001), cirrhotic background (p=0.002), age (p<0.001), blood loss (p<0.001) and liver resection for colorectal liver metastasis (p <0.001) were significantly associated with residual heterogeneity and hence postoperative morbidity. [Table 2]. Heterogeneity was significantly reduced with residual heterogeneity was non-significant with I2 of 39.99%, Q value 3.33, df 2, with the nonsignificant p-value. (p=0.189). Forest plot of multiple metaregression is included. [Figure 5]. Publication bias was nonsignificant with p=0.299. [Figure 6]. Diagnostic plots after multiple meta-regression analyses are shown in Supplement Figure 1.

**Table 2:**
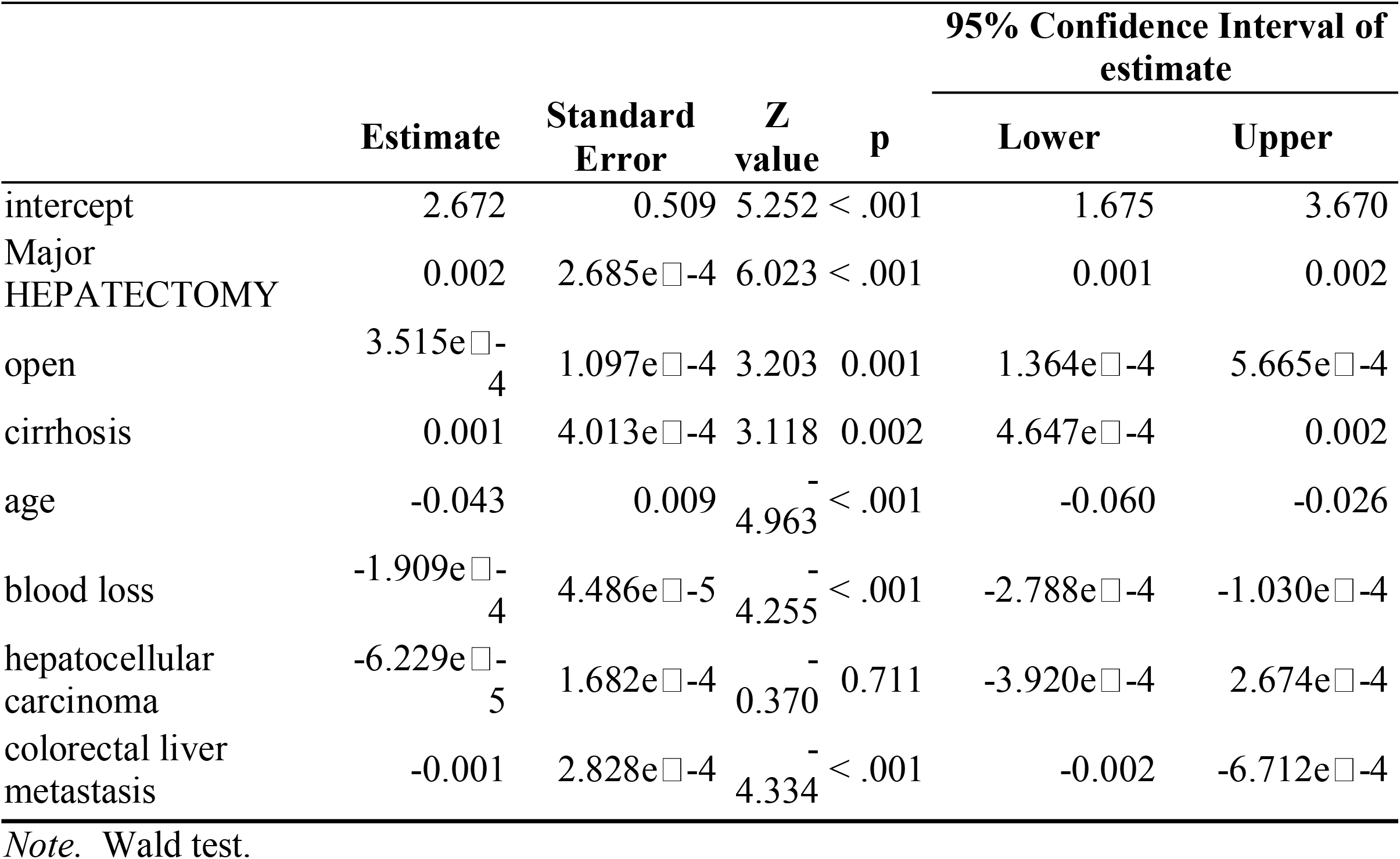
Multiple meta-regression of various factors.

**Figure 5.**
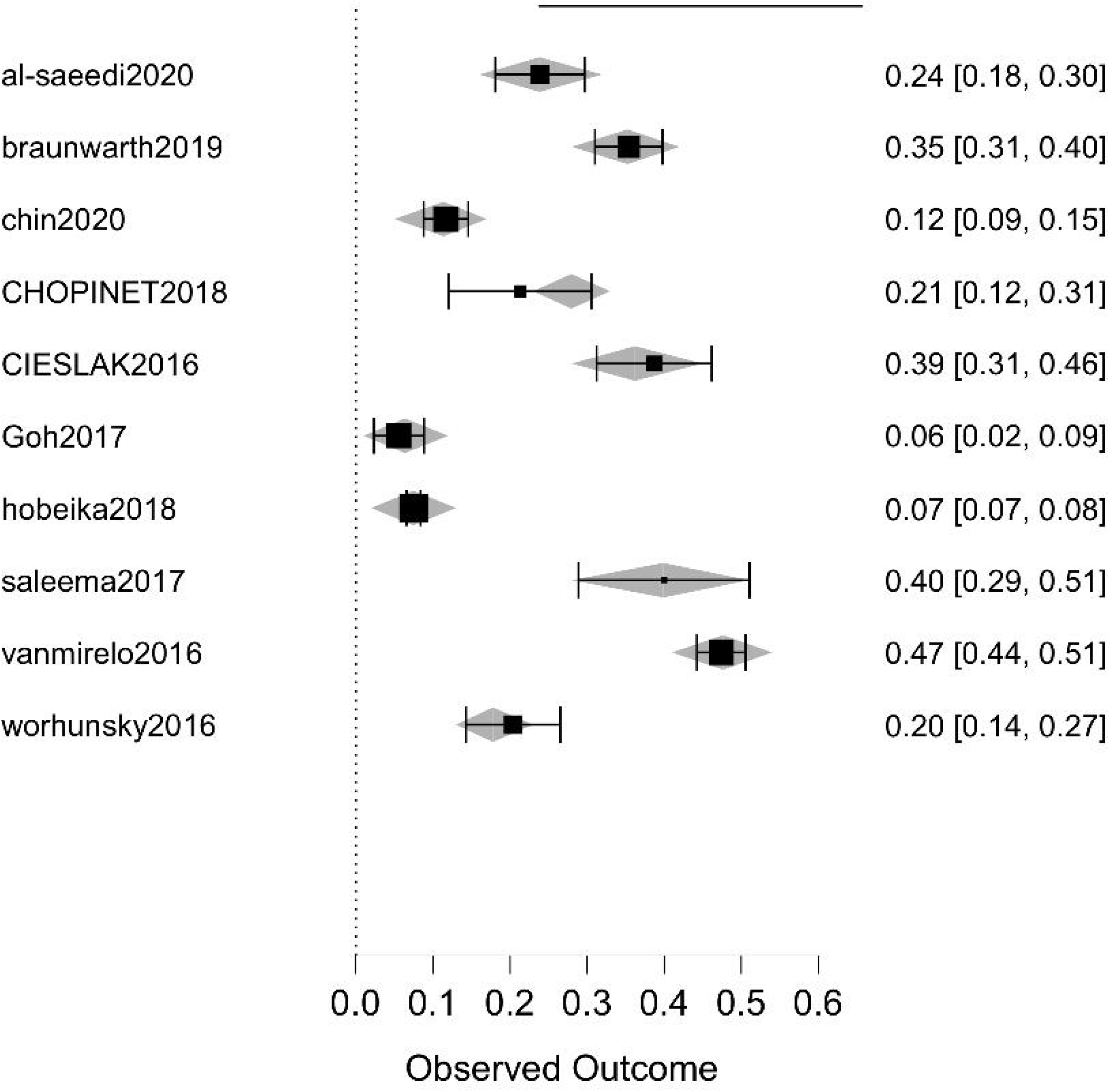
Forest plot of multiple meta-regression. (Observed outcomes fitted vs standardized outcomes calculated)

**Figure 6.**
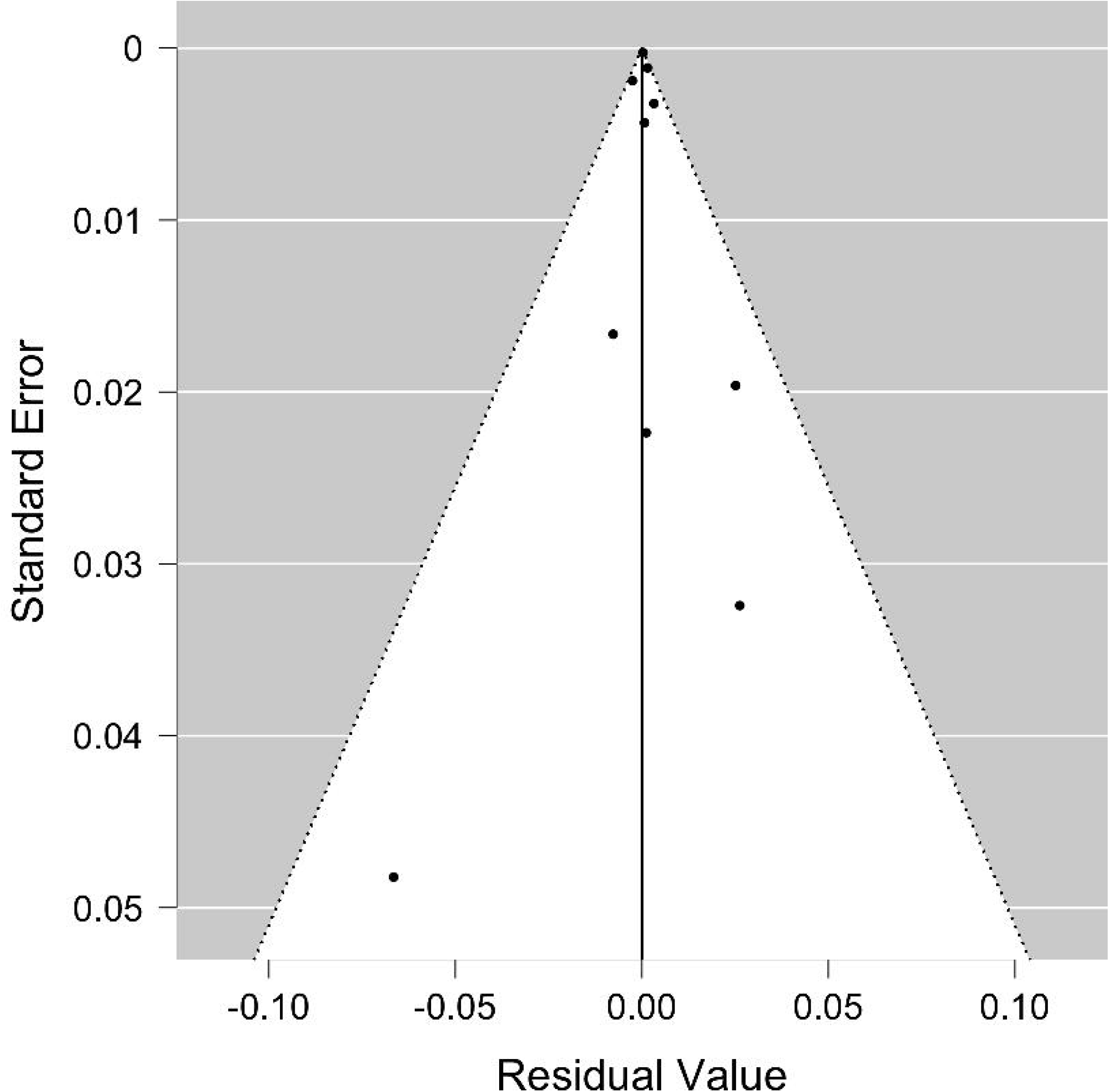
Funnel plot for studies mentioning all the factors in multiple metaregression.

## Discussion

Liver resection is a curative surgery for many benign and malignant disorders, with the most common indications are hepatocellular carcinoma and colorectal liver metastasis. Liver resection was historically associated with very high morbidity and mortality, which has now decreased significantly due to improved surgical and anaesthetic techniques and improved perioperative and critical care. Complications and hence morbidity rates though decreased but remain very high and range between 20-40%. [14-58]. There are also wide regional variations in indications of liver resections. In Asian countries, the most common indication is hepatocellular carcinoma [59] while in European countries most common indication is colorectal metastasis, which can also be the reason for variable mortality following liver resection.

Our aim in conducting this systemic review and prevalence meta-analysis to study weighted post-operative morbidity rates after liver resections. We also aimed to look at the heterogeneity of the analysis and publication bias. We also did metaregression analysis for various factors affecting mortality like Major hepatectomy, blood loss, age, open resections, a cirrhotic background of the liver, and etiologies for resections like hepatocellular carcinoma and colorectal metastasis in a study published in the last 5 year to look for recent trends.

As shown in figure 3, Weighted post-operative morbidity was 30.2% (95 % C.I. 24.8-\35.7%). However, the heterogeneity of the analysis was significantly high. On univariate metaregression, only major hepatectomy was significantly associated with heterogeneity and hence morbidity but residual heterogeneity was still significantly high. After multiple factor metaregression entering all above factors as covariates, residual heterogeneity was moderate and not significant with p=0.189 and I2 39.99%. Elimination of residual heterogeneity after metaregression suggested that the above factors were mainly responsible for variable outcomes across the centers.

On multiple factor metaregression major hepatectomy, age, open surgery, cirrhotic background, blood loss, and colorectal liver metastasis were associated with heterogeneity and morbidity. As shown in table 2 major hepatectomy, open surgery, and cirrhotic background were having positive metaregression coefficients and z value suggesting a positive relationship which means higher the number of major hepatectomies, open surgery and surgery on cirrhotic liver higher the chance of positive effect size and hence higher morbidities. Colorectal liver metastasis was having a negative metaregression coefficient and z value suggesting higher the number of liver resections for colorectal liver metastasis lower the effect size and hence morbidity.

Blood loss and age were having negative metaregression coefficients and z value suggestive of negative correlation or inverse relationship between effect size or morbidity and them. This might be due to patients with older age or more blood loss have higher post-operative mortality even before complications happen. Their positive correlation with 90 days postoperative mortality has been reported by us in a similar meta-analysis with metaregression analysis under review for publication and available as preprint online. [60]. Limitations of the meta-analysis were a large number of studies had to be excluded due to lack of complication data [61]. Also, we could not take into account center volume and surgeon’s experience. Another limitation was only 10 studies mentioned all the moderators however, the number of studies was adequate to conduct metaregression analysis.

However, to our knowledge, this is the first meta-analysis that evaluated weighted post-\operative morbidity rates and evaluated various factors responsible for heterogeneity and lack of significant residual heterogeneity after metaregression proved their effects on variable morbidity rates across the centers.

In conclusion, postoperative morbidity rates may vary according to centers based on various factors mentioned above. Major hepatectomy. In open surgery, the cirrhotic background may be associated with higher morbidities and liver resection colorectal liver metastasis may be associated with lower postoperative morbidity compared with other etiologies like hepatocellular carcinoma.

## Data Availability

can be made available on demand.

## Figure Legends

**Supplement Figure 1:**
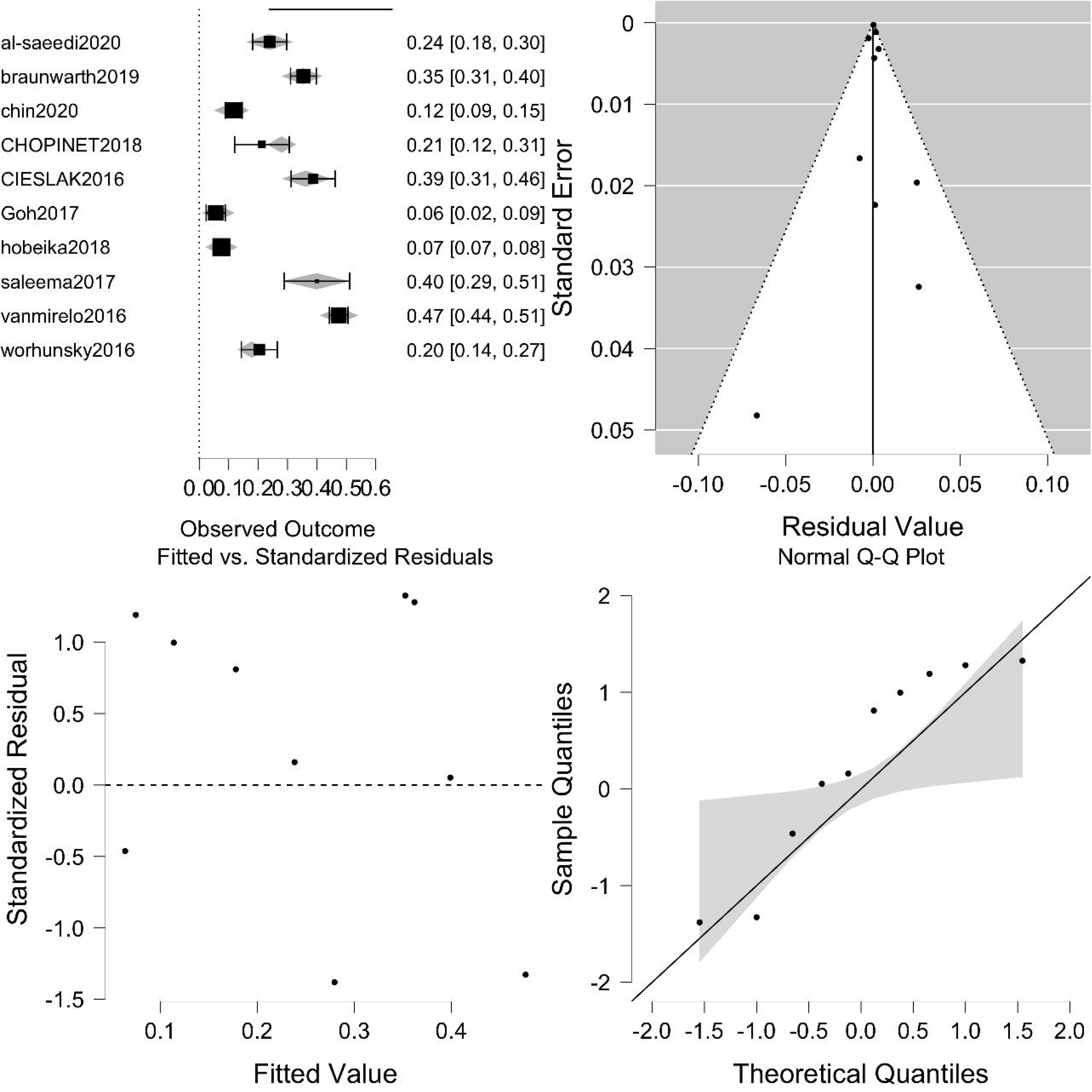
Diagnostic plots after multiple metaregression.

## Notes

Conflict of Interests: None

Funding disclosure: nothing to disclose.

### Competing Interest Statement

The authors have declared no competing interest.

### Funding Statement

no funding obtained.

### Author Declarations

systemic review and metaanalysis

## References

1. Wallace MC, Preen D, Jeffrey GP et al (2015) The evolving epidemiology of hepatocellular carcinoma: a global perspective. Expert Rev Gastroenterol Hepatol 9(6):765–779.

2. Menfredi S, Lepage C, Hatem C et al (2006) Epidemiology and management of liver metastases from colorectal cancer. Ann Surg. 244(2):254.

3. Gilg S, Sandstr Öm P, Rizell M et al (2018) The impact of post hepatectomy liver failure on mortality: a population-based study. Scand J Gastroenterol 25:1–5

4. Ray S, Mehta NN, Golhar A et al (2018) Post hepatectomy liver failure–A comprehensive review of current concepts and controversies.Ann Med Surg 34:4–10.

5. Kauffmann R, Fong Y (2014) Post-hepatectomy liver failure. Hepatobiliary Surg Nutr 3(5):238.

6. Chang CM, Yin WY, Su YC, et al. Preoperative risk score predicting 90-day mortality after liver resection in a population-based study. Medicine (Baltimore). 2014;93(12):e59.

7. Page M J, McKenzie J E, Bossuyt P M, et al. The PRISMA 2020 statement: an updated guideline for reporting systematic reviews BMJ 2021; 372 :71

8. Stroup DF, Berlin JA, Morton SC, et al. Meta-analysis of observational studies in epidemiology: a proposal for reporting. Meta-analysis Of Observational Studies in Epidemiology (MOOSE) group. JAMA. 2000;283(15):2008–2012.

9. Goossen K, Tenckhoff S, Probst P, et al. Optimal literature search for systematic reviews in surgery. Langenbecks Arch Surg. 2018;403(1):119–129.

10. Clavien PA, Barkun J, de Oliveira ML, et al. The Clavien-Dindo classification of surgical complications: five-year experience. Ann Surg. 2009;250(2):187–196.

11. Higgins JP, Thompson SG, Deeks JJ, Altman DG. Measuring inconsistency in meta-analyses. BMJ 2003; 327: 557–60.

12. Cochrane handbook for systematic reviews of interventions version 5.1.0 [updated March 2011]. Edited by Higgins JPT, Green S. [ http://www.cochrane-handbook.org].

13. Wells G, Shea B, O’Connell D, Peterson J, Welch V, Losos M, Tugwell P: The Newcastle-Ottawa Scale (NOS) for assessing the quality of nonrandomised studies in meta-analyses. In 2013.http://www.ohri.ca/programs/clinical_epidemiology/oxford.asp.

14. Al-Saeedi M, Ghamarnejad O, Khajeh E, et al. Pringle Maneuver in Extended Liver Resection: A propensity score analysis. Sci Rep. 2020;10(1):8847.

15. Al-Saif FA, Aldekhayel MK, Al-Alem F, Hassanain MM, Mattar RE, Alsharabi A. Comparison study between open and laparoscopic liver resection in a Saudi tertiary centre. Saudi Med J. 2019;40(5):452–457.

16. Al-Alem F, Mattar RE, Fadl OA, Alsharabi A, Al-Saif F, Hassanain M. Morbidity and mortality and predictors of outcome following hepatectomy at a Saudi tertiary care centre. Ann Saudi Med. 2016;36(6):414–421.

17. Amico EC, Alves JR, João SA, Guimarães PL, Medeiros JA, Barreto ÉJ. IMMEDIATE COMPLICATIONS AFTER 88 HEPATECTOMIES - BRAZILIAN CONSECUTIVE SERIES. Arq Bras Cir Dig. 2016 Jul-Sep;29(3):180–184.

18. Attili A, Sucandy I, Spence J, Bourdeau T, Ross S, Rosemurgy A. Outcomes of extended hepatectomy for hepatobiliary tumours. Initial experience from a non-university hepatobiliary centre. Am J Surg. 2020;219(1):106–109.

19. Braunwarth E, Primavesi F, Göbel G, et al. Is bile leakage after hepatic resection associated with impaired long-term survival?. Eur J Surg Oncol. 2019;45(6):1077–1083.

20. Chin KM, Koh YX, Syn N, et al. Early Prediction of Post-hepatectomy Liver Failure in Patients Undergoing Major Hepatectomy Using a PHLF Prognostic Nomogram. World J Surg. 2020;44(12):4197–4206.

21. Chopinet S, Grégoire E, Bollon E, et al. Short-term outcomes after major hepatic resection in patients with cirrhosis: a 75-case unicentric western experience. HPB (Oxford). 2019;21(3):352–360.

22. Cieslak KP, Bennink RJ, de Graaf W, et al. Measurement of liver function using hepatobiliary scintigraphy improves risk assessment in patients undergoing major liver resection. HPB (Oxford). 2016;18(9):773–780.

23. Fujii Y, Nanashima A, Hiyoshi M, Imamura N, Yano K, Hamada T. Risk factors for hepatic insufficiency after major hepatectomy in non-cirrhotic patients. Asian J Surg. 2019;42(1):251–255.

24. Garnier J, Faucher M, Marchese U, et al. Severe acute kidney injury following major liver resection without portal clamping: incidence, risk factors, and impact on short-term outcomes. HPB (Oxford). 2018;20(9):865–871.

25. Gelli M, Allard MA, Farges O, et al. Use of aspirin and bleeding-related complications after hepatic resection. Br J Surg. 2018;105(4):429–438.

26. Goh BK, Teo JY, Chan CY, et al. Evolution of laparoscopic liver resection at Singapore General Hospital: a nine-year experience of 195 consecutive resections. Singapore Med J. 2017;58(12):708–713.

27. Gray AD, Petrou G, Rastogi P, Begbie S. Elective hepatic resection is feasible and safe in a regional centre. ANZ J Surg. 2018 Mar;88(3):E147–E151.

28. Gupta AK, Kanhere HA, Maddern GJ, Trochsler MI. Liver resection in octogenarians: are the outcomes worth the risk? ANZ J Surg. 2018 Nov;88(11):E756–E760.

29. Halls MC, Cipriani F, Berardi G, et al. Conversion for Unfavourable Intraoperative Events Results in Significantly Worse Outcomes During Laparoscopic Liver Resection: Lessons Learned From a Multicentre Review of 2861 Cases. Ann Surg. 2018;268(6):1051–1057.

30. Higashi T, Hayashi H, Taki K, et al. Sarcopenia, but not visceral fat amount, is a risk factor of postoperative complications after major hepatectomy [published correction appears in Int J Clin Oncol. 2017 Oct;22(5):986-990]. Int J Clin Oncol. 2016;21(2):310–319.

31. Hobeika C, Fuks D, Cauchy F, et al. Impact of cirrhosis in patients undergoing laparoscopic liver resection in a nationwide multicentre survey. Br J Surg. 2020;107(3):268–277.

32. Lee KF, Chong C, Cheung S, et al. Robotic versus open hemi hepatectomy: a propensity score-matched study [published online ahead of print, 2020 Nov 13]. Surg Endosc. 2020;10.1007/s00464-020-07645-x.

33. Lemke M, Karanicolas PJ, Habashi R, et al. Elevated Lactate is Independently Associated with Adverse Outcomes Following Hepatectomy. World J Surg. 2017;41(12):3180–3188.

34. Levi Sandri GB, Colasanti M, Vennarecci G, et al. A 15-year experience of two hundred and twenty five consecutive right hepatectomies. Dig Liver Dis. 2017;49(1):50–56.

35. Lin S, Wu F, Wang L, et al. Surgical outcomes of hand-assisted laparoscopic liver resection vs. open liver resection: A retrospective propensity score-matched cohort study. Chin J Cancer Res. 2019;31(5):818–824.

36. Longbotham D, Young A, Nana G, et al. The impact of age on post-operative liver function following right hepatectomy: a retrospective, single centre experience. HPB (Oxford). 2020;22(1):151–160.

37. Donadon M, Fontana A, Palmisano A, et al. Individualized risk estimation for postoperative morbidity after hepatectomy: the Humanitas score. HPB (Oxford). 2017;19(10):910–918.

38. Ome Y, Hashida K, Yokota M, Nagahisa Y, Okabe M, Kawamoto K. The safety and efficacy of laparoscopic hepatectomy in obese patients. Asian J Surg. 2019;42(1):180–188.

39. Pietrasz D, Fuks D, Subar D, et al. Laparoscopic extended liver resection: are postoperative outcomes different?. Surg Endosc. 2018;32(12):4833–4840.

40. Ratti F, Cipriani F, Reineke R, et al. The clinical and biological impacts of the implementation of fast-track perioperative programs in complex liver resections: A propensity score-based analysis between the open and laparoscopic approaches. Surgery. 2018;164(3):395–403.

41. Ruzzenente A, Conci S, Ciangherotti A, et al. Impact of age on short-term outcomes of liver surgery: Lessons learned in 10-years’ experience in a tertiary referral hepato-pancreato-biliary centre. Medicine (Baltimore). 2017;96(20):e6955.

42. Begum S, Khan MR. Short-term outcomes after hepatic resection : perspective from a developing country. J Pak Med Assoc. 2017;67(8):1242–1247.

43. Urbani L, Colombatto P, Balestri R, et al. Techniques of parenchyma-sparing hepatectomy for the treatment of tumours involving the hepatocaval confluence: A reliable way to assure an adequate future liver remnant volume. Surgery. 2017;162(3):483–499.

44. van Mierlo KMC, Lodewick TM, Dhar DK, et al. Validation of the peak bilirubin criterion for outcome after partial hepatectomy. HPB (Oxford). 2016;18(10):806–812.

45. Viganò L, Torzilli G, Aldrighetti L, et al. Stratification of Major Hepatectomies According to Their Outcome: Analysis of 2212 Consecutive Open Resections in Patients Without Cirrhosis. Ann Surg. 2020;272(5):827–833.

46. Worhunsky DJ, Dua MM, Tran TB, et al. Laparoscopic hepatectomy in cirrhotics: safe if you adjust technique. Surg Endosc. 2016;30(10):4307–4314.

47. Yip VS, Dunne DF, Samuels S, et al. Adherence to early mobilisation: Key for successful enhanced recovery after liver resection. Eur J Surg Oncol. 2016;42(10):1561–1567.

48. Zaydfudim VM, Turrentine FE, Smolkin ME, Bauer TB, Adams RB, McMurry TL. The impact of cirrhosis and MELD score on postoperative morbidity and mortality among patients selected for liver resection. Am J Surg. 2020;220(3):682–686.

49. Xu LN, Yang B, Li GP, Gao DW. Assessment of complications after liver surgery: Two novel grading systems applied to patients undergoing hepatectomy. J Huazhong Univ Sci Technolog Med Sci. 2017;37(3):352–356.

50. Weinberg L, Mackley L, Ho A, et al. Impact of a goal directed fluid therapy algorithm on postoperative morbidity in patients undergoing open right hepatectomy: a single centre retrospective observational study. BMC Anesthesiol. 2019;19(1):135.

51. Ueno M, Kawai M, Hayami S, et al. Partial clamping of the infrahepatic inferior vena cava for blood loss reduction during anatomic liver resection: A prospective, randomized, controlled trial. Surgery. 2017;161(6):1502–1513.

52. Uchida H, Iwashita Y, Tada K, et al. Laparoscopic liver resection in cirrhotic patients with specific reference to a difficulty scoring system. Langenbecks Arch Surg. 2018;403(3):371–377.

53. Tran TB, Worhunsky DJ, Spain DA, et al. The significance of underlying cardiac comorbidity on major adverse cardiac events after major liver resection. HPB (Oxford). 2016;18(9):742–747.

54. Snyder RA, Ewing JA, Parikh AA. Preoperative Portal Vein Embolization Is Not Associated with Increased Postoperative Complications After Major Hepatectomy: a Study of the National Surgical Quality Improvement Database. J Gastrointest Surg. 2020;24(7):1561–1570.

55. Shwaartz C, Fields AC, Aalberg JJ, Divino CM. Role of Drain Placement in Major Hepatectomy: A NSQIP Analysis of Procedure-Targeted Hepatectomy Cases. World J Surg. 2017 Apr;41(4):1110–1118.

56. Ross SW, Seshadri R, Walters AL, et al. Mortality in hepatectomy: Model for End-Stage Liver Disease as a predictor of death using the National Surgical Quality Improvement Program database. Surgery. 2016;159(3):777–792.

57. Le Roux F, Rebibo L, Cosse C, et al. Benefits of Laparoscopic Approach for Resection of Liver Tumours in Cirrhotic Patients. J Laparoendosc Adv Surg Tech A. 2018;28(5):553–561.

58. Hughes MJ, Chong J, Harrison E, Wigmore S. Short-term outcomes after liver resection for malignant and benign disease in the age of ERAS. HPB (Oxford). 2016;18(2):177–182.

59. Chang CM, Yin WY, Su YC, et al. Preoperative risk score predicting 90-day mortality after liver resection in a population-based study. Medicine (Baltimore). 2014;93(12):e59.

60. Bhavin Vasavada, Hardik Patel. Recent trends in postoperative mortality after liver resection- A systemic review and metanalysis of studies published in last 5 years and metaregression of various factors affecting 90 days mortality. medRxiv 2021.03.26.21254407; doi:https://doi.org/10.1101/2021.03.26.21254407.

61. Filmann N, Walter D, Schadde E, et al. Mortality after liver surgery in Germany. Br J Surg. 2019;106(11):1523–1529.

